# The Cost of Social Care Need in People with Multimorbidity: A Population-Based Cohort Study of 5.7 million Individuals

**DOI:** 10.1101/2025.10.29.25339063

**Authors:** Tassella Isaac, Aman Jat, Lucy Smith, Yousef Yousef, Rishika Vashishtha, Mehedi Hasan, Hajira Dambha-Miller

## Abstract

**Background:** Multimorbidity (the coexistence of two or more long-term conditions) is increasingly prevalent, particularly among older adults, and is a major driver of healthcare expenditure. However, healthcare costs reflect only part of the overall burden. Many people with multimorbidity also depend on social care services for personal support, mobility assistance, and social engagement to preserve independence and quality of life. Despite its significance for individuals, families, and the health system, the economic impact of social care needs related to multimorbidity has not been quantified at a population level in England.

**Methods:** We conducted a retrospective, population-based cohort study of adults (≥18 years) with multimorbidity using data from the CPRD Gold and Aurum datasets covering 1987–2020. Social care needs were identified through coded indicators across three domains (personal care, mobility, and social interaction) and valued using 2019–2020 national unit costs from the Personal Social Services Research Unit (PSSRU). Adjusted per-capita costs were estimated via direct standardisation by age, sex, ethnicity, and socioeconomic deprivation.

**Results:** The cohort comprised 5,771,603 adults with multimorbidity (mean age 56.5 years; 55.1% female). Among them, 800,309 individuals (13.9%) had at least one recorded social care need. Personal care accounted for the majority of costs (£3.66 billion; 84.3% of total), affecting 7.3% of the cohort with a mean annual expenditure of £8,736 per user. Mobility needs represented £336.8 million (7.8%) and social interaction needs £346 million (8.0%), with mean annual costs of £1,426 and £709 per user, respectively. Individuals with multiple needs (6.3% of the cohort) incurred mean annual costs of £9,076. Social care needs and costs were higher among older adults, women, and those living in more deprived areas.

**Conclusion:** This study provides the first population-level estimate of social care costs associated with multimorbidity in England. Social care represents an important yet often overlooked component of the broader economic burden of multimorbidity. Costs are predominantly concentrated in personal care and vary systematically by demographic and socioeconomic factors, underscoring potential inequities in service access or provision. Integrating social care within multimorbidity strategies is essential to address the full scale of its societal impact.

## Background

Multimorbidity, defined as the co-occurrence of two or more long-term health conditions within an individual, is increasingly prevalent in the United Kingdom and globally. In England, approximately two-thirds of individuals aged 65 years and over are projected to live with multimorbidity by 2035, with a large proportion managing four or more conditions [1]. The consequences of multimorbidity include poorer quality of life, increased frailty, polypharmacy, and significantly greater use of healthcare services. A study [2] demonstrated that individuals with multimorbidity accounted for the majority of general practice consultations and hospital admissions, particularly in deprived areas. While healthcare costs associated with multimorbidity have been extensively studied, the economic implications for social care services remain poorly quantified.

Social care needs consist of non-medical support that enables people to maintain independence and quality of life. Traditionally, this has focused on activities of daily living (ADL), such as personal hygiene, eating, and dressing. The concept of instrumental activities of daily living (IADL) broadened this scope to include tasks essential for independent living, such as managing medications and shopping [3]. The Care Act 2014 in England further formalised eligibility, using a broader definition of social care need [4]. Social care need represents a vital aspect of the overall burden of multimorbidity, encompassing personal care, mobility, and social interaction. These needs may be met through formal services, unpaid carers, or remain unmet, yet they carry significant economic and human costs. As local government funding for adult social care in England has declined in real terms over the past decade, reliance on unpaid carers and community services has grown [5,6]. The current literature has largely prioritised medical utilisation costs, overshadowing the contribution of social care need to overall care expenditure.

Recent research has begun to address this imbalance. One study [7] conducted an ecological analysis in England and found that areas with higher multimorbidity prevalence among older adults had significantly greater local authority spending on adult social care. However, this relied on area-level correlations and did not examine person-level patterns or adjust for individual characteristics. A further cohort study [8] analysed multimorbidity patterns in individuals with Alzheimer’s disease, while many other cost studies rely on survey data or evidence from non-UK settings, limiting policy relevance [9]. There is therefore a need for population-level, individual-based analysis of social care need and its associated costs in people with multimorbidity using routinely collected data in the UK. The current evidence base remains focused on healthcare utilisation and costs, with little attention to how multimorbidity translates into demand for, and cost of, social care needs. Recent systematic reviews have highlighted this gap, noting that the social care need costs of multimorbidity remain largely unquantified [9]. This study addresses this gap by quantifying the costs of social care need in individuals with multimorbidity in England, using routinely collected, population-based data. Specifically, we estimate the annual and lifetime costs of social care need and identify sociodemographic and clinical predictors of higher costs.

## Methods

### Data source and study design

We conducted a retrospective, population-based cohort study using routinely collected, anonymised electronic health records from England. We used data from the Clinical Practice Research Datalink (CPRD) Gold and AURUM databases [10–13], coded using the READ, ICD-10 and SNOMED systems, and linked to Hospital Episode Statistics (HES) and Office for National Statistics (ONS) records [14,15].

CPRD is a government-run data service providing longitudinal UK medical records from primary care [16]. It contains information from over 65 million patients across the UK, including over 19 million currently registered patients [14]. The database is nationally representative of the UK population by age, sex, ethnicity, and socioeconomic status, as measured by the English Index of Multiple Deprivation (IMD) at the level of the Lower Super Output Area of residence [17]. Geographic region was defined based on the postcode of the patient’s registered general practice and mapped to one of the standard NHS administrative regions in England. CPRD includes coded data on clinical events, medical diagnoses, referrals to hospital and other specialist care settings, prescriptions, and related patient information. These data were linked using unique patient identifiers. CPRD is widely used for public health research, including studies on the burden of disease, risk factors, outcomes, and patterns of healthcare utilisation. For this study, we included adults aged 18 years and older with multimorbidity, defined as two or more long-term conditions, and with records between 1 January 1987 and 31 December 2020 who met the data-quality requirements. (Supplementary Table 1)

Social care need information was coded using mutually exclusive variables representing different types of support. This structure was based on a previously established multidimensional framework for identifying social care needs among people with multimorbidity [18]. Three domains from this framework were used in the present study: personal care needs, mobility needs, and social interaction need. Social care need costs were extracted from the 2019–2020 Personal Social Services Research Unit (PSSRU) cost compendium and mapped to CPRD-recorded social care needs according to service type, age group, and condition category. Where possible, local authority-specific unit costs were used as baseline estimates. Further details on the definitions of social care need and the costing methodology are provided below.

### Definition of Multimorbidity

Multimorbidity was defined as having two or more long-term conditions (LTCs) [19]. The list of conditions was based on a consensus-derived set of 59 LTCs developed by UK researchers relevant to multimorbidity research [20]. Of these, 55 conditions were defined in CPRD using validated code lists, conditions with low counts or clinical overlap were grouped into broader categories. The co-existence of any two or more of these conditions constituted multimorbidity. The conditions covered a wide range of physical and mental health disorders (e.g., diabetes, asthma, coronary heart disease), mental health conditions (e.g., schizophrenia, depression), sensory impairments (e.g., hearing and vision loss), disabilities (including learning disabilities), chronic pain syndromes, and alcohol or substance misuse. For each individual, the date of diagnoses of the second condition was defined as the index date [21]. Full condition definitions and corresponding SNOMED and ICD-10 codes are provided in Supplementary Tables 2 and 3.

### Definition of Social Care Need

Social care needs were identified using a previously established framework that defined eight social care need domains relevant to people with multimorbidity. Variables were extracted from electronic health records and mapped to CPRD primary care data using over 100 mutually exclusive codes. For this study, three domains of social care need were included, selected according to the scope of analysis and the availability of public cost data. These were Personal Care Needs, Mobility Needs, and Social Interaction Needs:

- Personal Care Needs referred to difficulties with activities of daily living, including ambulation, feeding, dressing, personal hygiene, continence and toileting.
- Mobility needs encompassed difficulties with physical mobility requiring aids or professional support, such as the use of wheelchairs or other mobility devices, or assistance from care staff.
- Social Interaction Needs captured limitations in an individual’s ability to socially connect and participate in social or community activities. This included relationships with family and friends, verbal and non-verbal communication, participation in hobbies or leisure activities, community activities, community engagement, ability to travel, and receipt of professional or voluntary support to address loneliness or social isolation.

A full list of social care need variables and corresponding codes are provided in the Supplementary Tables 4 and 5. Multiple needs were defined as having two or more domains positive within the same person-year.

### Costs Calculation

Costs were estimated in three stages: unit cost mapping, individual-level assignment, and aggregation to annual equivalents. Unit costs for social care needs were obtained from the 2019–2020 Personal Social Services Research Unit (PSSRU) cost compendium and expressed in 2019–2020 British pounds [22]. The 2019–2020 PSSRU cost compendium year was selected as it corresponds to the final year of the study period (1 January 1987–31 December 2020), ensuring alignment between the costing year and the observation window. Each social care need variable recorded in CPRD was mapped to the most appropriate PSSRU unit cost according to provider type, setting, client group, age band, and unit type. A deterministic, rule-based “closest-match” algorithm was implemented to assign costs at the individual level. For each combination of setting, client group, and age band, each cost record received an age-match score and a client-group score values as 2 for an exact match, 1 for a compatible match (e.g., age 18-64 matched to “All adults”, or age ≥65 matched to “65+” or “All adults”), and 0.5 when unspecified. A setting score of 1 was given when the cost setting overlapped the CPRD setting. The total score was computed as 1.5 x age + 1.5 x client +1 x setting, and the row with the highest total score was selected by source code. Where relevant, regional adjustments were applied (+14% for private nursing homes and +18% for private residential care) based on the specifications provided in the PSSRU cost sheet to account for London uplifts. Costs were expressed as annual equivalents per person-year. Per-hour tariffs (for example, home care or peer support) were converted using default weekly assumptions, and per-episode costs (for example, reablement) were applied once per year.

Personal-care costs were assigned hierarchically to ensure one package per person-year. Residential tariffs (nursing or residential care) were prioritised over community hourly tariffs and supported-living or learning-disability lines were applied when relevant. Mobility costs included wheelchair and equipment provision, home adaptations, and reablement services. Social-interaction costs included day care, social prescribing, and peer support, each matched to age group and service type.

To avoid double counting, residential personal-care tariffs took precedence over community or mobility costs, and only one residential category was assigned per person-year. Sensitivity analyses tested alternative assumptions for home-care hours, inclusion of day services, and variation in unit prices. Full details of cost mappings, scoring logic, uplifts, and sensitivity settings are provided in the Supplementary Tables 7-8.

### Outcomes

The main outcome was annual social care needs cost per person-year. Domain-specific and total annual costs were derived for each person. Individuals without any recorded social-care use were assigned a cost of £0 for that domain and year. Secondary outcomes were per-capita adjusted annual costs, estimated by age group, sex, ethnicity, and Index of Multiple Deprivation (IMD) quintile. All outcomes were derived for adults aged 18 years and older during the study window (1 January 1987 to 31 December 2020). Denominators for per-capita estimates included all individuals, including those with zero costs or events.

### Covariates

Covariates included age, sex, ethnicity, Index of Multiple Deprivation (IMD) and region of residence. Age was grouped into five categories: 18–44, 45–64, 65–74, 75–84, and 85 years or older. Sex was coded as Male or Female, with missing or indeterminate values coded as Unknown. Ethnicity was derived from linked HES data and mapped into five categories (White, Asian, Black, Mixed, and Other), with missing values recorded as Unknown. Socioeconomic status was measured using the English Index of Multiple Deprivation (IMD) linked to each patient’s Lower Super Output Area of residence. Patient-level IMD was mapped to quintiles Q1 (least deprived) to Q5 (most deprived).

When patient-level IMD was missing or invalid, practice-level IMD was used as a fallback, if both were missing, the composite IMD field remained Unknown. Region of residence was derived from the practice region information. A binary flag identified London residents for analyses requiring regional uplift adjustments.

Missing data were minimal (≤1.3%) for all variables except ethnicity (16%), which were coded as ‘Unknown’ and carried forward to the analysis.

### Statistical analysis

Each social care need domain (Personal Care, Mobility, and Social Interaction) was represented in CPRD by a set of mutually exclusive indicators. A domain was considered present for an individual if any subcategory equalled 1, or if the total count of indicators was greater than zero or if the main domain indicator was recorded as 1. Based on the three domain-level flags, a mutually exclusive exposure variable was created:

1. None: none of the social care needs present
2. Personal Care only, Mobility only, or Social Interaction only: exactly one of the social care needs present and
3. Multiple: two or more social care needs present within the same person-year.

Baseline descriptive analyses were calculated with consistent denominators. Categorical variables were summarised as counts and column percentages by exposure group, with Unknown and Missing categories reported. Continuous variables were summarised as mean (standard deviation) and median (interquartile range).

We also calculated costs specific to each social care need for Personal Care, Mobility, Social Interaction, and Multiple Needs (≥2 domains with cost > £0). For each domain, we reported the number of people with non-zero annual spend, prevalence as a percentage of the total cohort, mean annual cost among spenders, total cohort spend, and spend per 100,000 adults, calculated as (total spend ÷ cohort size) × 100,000, equivalent to prevalence × mean × 100,000. These metrics are presented in the results tables to summarise both the distribution and relative contribution of each social care need domain to overall social care expenditure.

The share of total spends (%) was calculated across Personal Care, Mobility, and Social Interaction totals (summing to 100%). The Multiple needs group was reported descriptively and not added to the domain totals to prevent double counting. We estimated adjusted per-capital annual costs for each social care need (Personal Care, Mobility, and Social Interaction) across age group, sex, ethnicity, and Index of Multiple Deprivation (IMD) quintile. Costs were computed per person per year, including zero values for non-users, to reflect population-average spending within each subgroup. Direct standardisation was used to derive population-level means for subgroup comparisons, as recommended by the Healthcare Cost and Utilization Project [23,24]. For a target variable (e.g., age group), data was divided into small cells defined by the cross-classification of other covariates (for age: sex, ethnicity, IMD, and region). Within each target level (L) and cell (c), we calculated the cell-specific per-capita mean cost and variance using all individuals in that intersection (including zeros).

The adjusted mean cost for level L was estimated as a weighted average of cell means: Adjusted mean (μL) = Σ (wc × mean cost in cell c for level L) where wc represents the proportion of the overall cohort in that cell (the reference weight). This approach standardises subgroup means to the full population and avoids confounding by compositional differences. Uncertainty was quantified using the variance of a weighted mean (delta method): Var(μL) = Σ [ (wc^2^ × variance in cell c for level L) / (number of individuals in that cell)]. The standard error was the square root of this variance: SE(μL) =√Var(μL) and the 95% confidence interval was given by: μL ± 1.96 × SE(μL).

Costs for non-users were set to £0 so that per-capita estimates reflected population means. Denominators for each subgroup included all adults aged ≥ 18 years.

### Missingness

Unknown categories were retained in both standardisation cells and tabular outputs to maintain transparency. Where a target-by-cell combination contained a single observation, its variance contribution was treated as zero and down-weighted automatically in the weighted mean.

### Software

All analyses were conducted in R (version 4.3.2) using the data-table package for aggregation. No regression or resampling was required, the approach relied on grouped summaries and analytic variance, providing stable and scalable inference for the large dataset.

## Results

### Patient characteristics

Our final study population consisted of 5 771 603 adults aged 18 years and over, recorded between 1987-2020 (figure 1). Of these, 4 971 294 (86.1%) had no recorded social care need, 75 574 (1.3%) had personal care needs only, 161 361 (2.8%) had mobility needs only, 201 812 (3.5%) had social interaction needs only, and 361 562 (6.3%) had multiple social care needs. Overlap between Personal Care needs, Mobility needs and Social Interaction needs is shown in Supplementary Table 9. The most common overlap occurred between Personal care needs and Social interaction needs (n = 286 579).

**Figure 1.**
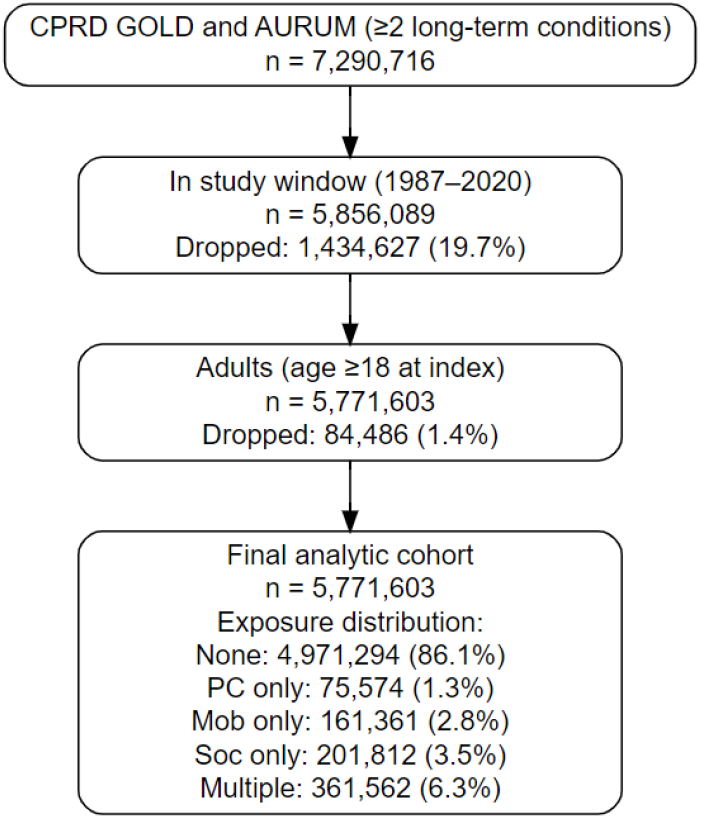
Flow of study population selection from CPRD Gold and AURUM (1987-2020) linked to Hospital Episode Statistics (HES) and Office for National Statistics (ONS). After initial extraction (n = 7 290 716), records were restricted to individuals with an index date between January 1, 1987, and December 31, 2020 (n = 5 856 089). Adults aged ≥18 years at index were retained, yielding the final study population of 5 771 603 individuals with multimorbidity (defined as ≥2 long-term conditions). Exposure distribution by social care need was none (4 971 294; 86.1%), personal care (75 574; 1.3%), mobility (161 361; 2.8%), social interaction (201 812; 3.5%), and multiple needs (361 562; 6.3%).

Baseline characteristics by exposure group are presented in Table 1. The mean age in the overall cohort was 56.5 years (SD 17.5), increasing in the social care need groups (66.1 years among those with mobility needs and 63.6 years among those with multiple needs). The median age was 58 years overall and 68 years among those with mobility needs. Women constituted 55.1% of the cohort, with slightly higher proportions in the mobility (60.1%) and social interaction (59.8%) groups compared with men. Deprivation was evenly distributed across quintiles, with about one-fifth of the population in each IMD quintile. Individuals with mobility needs were more deprived, with 23.2% living in the most deprived quintile (IMD 5) compared with 20.9% overall. The study population was predominantly White (75.8%), with 3.3% Asian, 2.1% Black, 0.5% Mixed, and 1.1% Other ethnic groups, ethnicity was Unknown for 16%. Regional representation covered all major NHS regions in England, with the largest contributions from the North-West (20.3%), South Central (19.8%), and South-West (14.7%) regions. London accounted for 13.5% of the study population. Across the cohort, the most frequently recorded long-term conditions were depression (32.7%) and anxiety (30.7%), followed by asthma (26.2%), arrhythmia (26.0%), and diabetes (22.9%). Thyroid disease (14.0%), heart failure (14.3%), Chronic Obstructive Pulmonary Disease (COPD) (3.2%), osteoarthritis (2.2%), and cancer (1.4%) completed the top ten by overall prevalence. People with multiple social care needs had consistently higher prevalences than those with no needs for most conditions, for example anxiety (47.6% vs 28.7%), depression (28.6% vs 33.0% in None; still high in Multiple given competing multimorbidity burden), diabetes (32.3% vs 21.7%), arrhythmia (27.1% vs 25.8%), asthma (32.4% vs 25.6%), thyroid disease (18.2% vs 13.3%), COPD (2.3% vs 3.2% overall but 3.5% in Mobility), and heart failure (26.1% vs 12.8%). Patterns differed by need: Mobility-only showed the highest proportions for diabetes (38.7%), heart failure (30.7%), anxiety (54.5%), and arrhythmia (25.5%). Social-interaction-only had the highest depression (41.3%) and relatively high arrhythmia (31.7%). Personal-care-only had anxiety (39.3%), asthma (31.4%) and diabetes (31.2%).

**Table 1.** Characteristics of adults with multimorbidity in England, by social care need exposure (1987-2020). Characteristics of 5,771 603 adults aged ≥18 years with multimorbidity identified from the Clinical Practice Research Datalink (CPRD) Gold and Aurum datasets linked to Hospital Episode Statistics and Office for National Statistics mortality data. Values are counts (n) and column percentages unless otherwise stated; unknown and missing values are reported. Exposure groups were defined as: none (n = 4 971 294), personal-care needs only (n = 75 574), mobility needs only (n = 161 361), social-interaction needs only (n = 201 812), and multiple needs (two or more domains positive within the same person-year; n = 361 562). Mean (SD) and median (IQR) are shown for continuous variables. IMD = Index of Multiple Deprivation.

| Variable | Level | Overall | None | Multiple needs | Personal Care needs | Mobility needs | Social interaction needs |
| --- | --- | --- | --- | --- | --- | --- | --- |
| **Socio-demographic** |  |  |  |  |  |  |  |
| **Age at index (years)** | Mean (SD) | 56.5 (17.5) | 55.5 (17.6) | 63.6 (15.6) | 59.5 (16.4) | 66.1 (14.4) | 57.8 (17.7) |
| **Age at index (years)** | Median [IQR] | 58.0 [44.0–70.0] | 57.0 [43.0–69.0] | 66.0 [55.0–75.0] | 61.0 [49.0–71.0] | 68.0 [57.0–77.0] | 59.0 [45.0–72.0] |
| **Sex** | Male | 2,593,497 (44.9) | 2,248,639 (45.2) | 163,931 (45.3) | 35,410 (46.9) | 64,355 (39.9) | 81,162 (40.2) |
|  | Female | 3,178,043 (55.1) | 2,722,598 (54.8) | 197,630 (54.7) | 40,163 (53.1) | 97,006 (60.1) | 120,646 (59.8) |
|  | Missing | 63 (0.0) | 57 (0.0) | 1 (0.0) | 1 (0.0) | 0 (0.0) | 4 (0.0) |
| **IMD** | IMD 1 (least) | 1,106,166 (19.2) | 952,716 (19.2) | 76,026 (21.0) | 14,577 (19.3) | 28,291 (17.5) | 34,556 (17.1) |
|  | IMD 2 | 1,157,946 (20.1) | 996,555 (20.0) | 77,279 (21.4) | 16,059 (21.2) | 31,462 (19.5) | 36,591 (18.1) |
|  | IMD 3 | 1,131,140 (19.6) | 971,615 (19.5) | 72,256 (20.0) | 16,581 (21.9) | 31,543 (19.5) | 39,145 (19.4) |
|  | IMD 4 | 1,171,277 (20.3) | 1,009,742 (20.3) | 68,247 (18.9) | 14,747 (19.5) | 32,570 (20.2) | 45,971 (22.8) |
|  | IMD 5 (most) | 1,205,074 (20.9) | 1,040,666 (20.9) | 67,754 (18.7) | 13,610 (18.0) | 37,495 (23.2) | 45,549 (22.6) |
| **Ethnicity** | Asian | 188,992 (3.3) | 157,856 (3.2) | 12,550 (3.5) | 2,459 (3.3) | 5,217 (3.2) | 10,910 (5.4) |
|  | Black | 121,586 (2.1) | 102,364 (2.1) | 5,751 (1.6) | 1,208 (1.6) | 2,982 (1.8) | 9,281 (4.6) |
|  | Mixed | 27,691 (0.5) | 24,368 (0.5) | 1,139 (0.3) | 282 (0.4) | 469 (0.3) | 1,433 (0.7) |
|  | Other | 63,954 (1.1) | 55,115 (1.1) | 3,210 (0.9) | 723 (1.0) | 1,354 (0.8) | 3,552 (1.8) |
|  | White | 4,376,824 (75.8) | 3,744,992 (75.3) | 290,601 (80.4) | 61,281 (81.1) | 128,811 (79.8) | 151,139 (74.9) |
|  | Unknown | 922,948 (16.0) | 821,600 (16.5) | 46,486 (12.9) | 9,274 (12.3) | 21,691 (13.4) | 23,897 (11.8) |
|  | Missing | 69,608 (1.2) | 64,999 (1.3) | 1,825 (0.5) | 347 (0.5) | 837 (0.5) | 1,600 (0.8) |
| **Region** | East Midlands | 153,561 (2.7) | 137,452 (2.8) | 7,201 (2.0) | 2,210 (2.9) | 3,478 (2.2) | 3,220 (1.6) |
|  | East of England | 312,758 (5.4) | 271,615 (5.5) | 18,998 (5.3) | 5,639 (7.5) | 8,570 (5.3) | 7,936 (3.9) |
|  | London | 778,299 (13.5) | 677,440 (13.6) | 43,019 (11.9) | 10,968 (14.5) | 23,839 (14.8) | 23,033 (11.4) |
|  | North-East | 216,340 (3.7) | 184,935 (3.7) | 14,871 (4.1) | 2,243 (3.0) | 5,326 (3.3) | 8,965 (4.4) |
|  | North-West | 1,173,785 (20.3) | 1,003,023 (20.2) | 79,047 (21.9) | 14,511 (19.2) | 40,009 (24.8) | 37,195 (18.4) |
|  | South Central | 1,143,874 (19.8) | 998,147 (20.1) | 68,126 (18.8) | 15,880 (21.0) | 26,603 (16.5) | 35,118 (17.4) |
|  | South-West | 849,177 (14.7) | 709,667 (14.3) | 54,555 (15.1) | 9,899 (13.1) | 22,184 (13.7) | 52,872 (26.2) |
|  | West Midlands | 906,776 (15.7) | 783,349 (15.8) | 61,781 (17.1) | 9,384 (12.4) | 23,993 (14.9) | 28,269 (14.0) |
|  | Yorkshire & The Humber | 233,926 (4.1) | 202,829 (4.1) | 13,858 (3.8) | 4,822 (6.4) | 7,243 (4.5) | 5,174 (2.6) |
|  | Missing | 3,107 (0.1) | 2,837 (0.1) | 106 (0.0) | 18 (0.0) | 116 (0.1) | 30 (0.0) |
| **Clinical variables** |  |  |  |  |  |  |  |
| **Long-term Conditions** | Depression | 2, 387, 124 (32.7%) | 2,135,960 (33.0%) | 111,331 (28.6%) | 16,383 (27.8%) | 44,091 (25.2%) | 79,359 (41.3%) |
|  | Anxiety | 2, 238, 026 (30.7%) | 1,861,907 (28.7%) | 185,103 (47.6%) | 23,127 (39.3%) | 95,255 (54.5%) | 72,634 (37.8%) |
|  | Asthma | 1,909,517 (26.2%) | 1,657,184 (25.6%) | 126,047 (32.4%) | 18,490 (31.4%) | 57,395 (32.8%) | 50,401 (26.2%) |
|  | Arrhythmia | 1,898,532 (26.0%) | 1,672,943 (25.8%) | 105,288 (27.1%) | 14,800 (25.1%) | 44,613 (25.5%) | 60,888 (31.7%) |
|  | Diabetes | 1,667,556 (22.9%) | 1,408,358 (21.7%) | 125,556 (32.3%) | 18,334 (31.2%) | 67,625 (38.7%) | 47,683 (24.8%) |
|  | Heart failure | 1,039,449 (14.3%) | 831,946 (12.8%) | 101,324 (26.1%) | 14,178 (24.1%) | 53,597 (30.7%) | 38,404 (20.0%) |
|  | Thyroid disease | 1,017,480 (14.0%) | 860,689 (13.3%) | 70,918 (18.2%) | 10,492 (17.8%) | 39,647 (22.7%) | 35,734 (18.6%) |
|  | COPD | 232,145 (3.2%) | 208,672 (3.2%) | 9,018 (2.3%) | 1,467 (2.5%) | 6,087 (3.5%) | 6,901 (3.6%) |
|  | Osteoarthritis | 162,542 (2.2%) | 136,640 (2.1%) | 13,041 (3.4%) | 1,918 (3.3%) | 6,511 (3.7%) | 4,432 (2.3%) |
|  | Cancer | 104,771 (1.4%) | 92,101 (1.4%) | 4,857 (1.2%) | 816 (1.4%) | 2,911 (1.7%) | 4,086 (2.1%) |

Annual social care need costs by domain are summarised in Table 2. Across the full adult study population, personal care needs accounted for the largest share of total expenditure, representing £3.66 billion (84.3%) of all recorded costs. This corresponds to 418 578 recipients (7.3% of the cohort) with a mean annual cost of £8 736 per user, equivalent to £63.4 million per 100, 000 adults. Mobility needs contributed £336.8 million (7.8%), covering 236 291 individuals (4.1% of the cohort) with a mean annual cost of £1 426 per user and £5.8 million per 100, 000 adults. Social interaction needs accounted for £346 million (8%) across 488 282 users (8.5%), with a mean annual cost of £709 per user and £6 million per 100 000 adults. Among those with multiple needs, the average annual cost reached £9, 076, with a total of £3.11 billion, however, these totals are reported for context only, as they are not additive with single need categories.

**Table 2.**
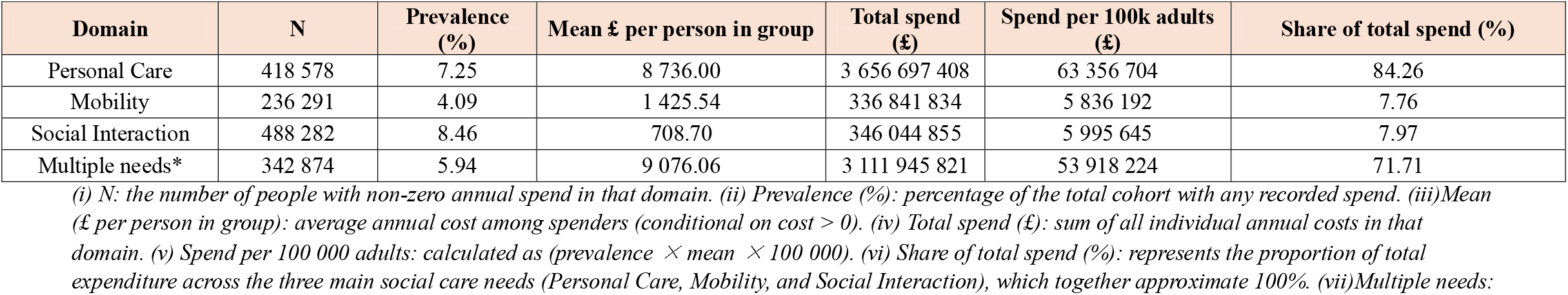
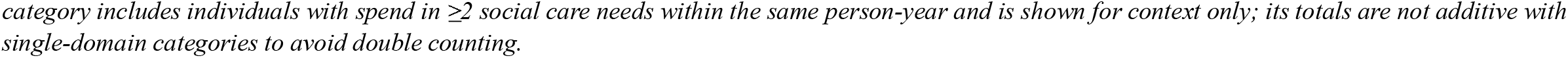
Annual social-care costs by domain among adults with multimorbidity in England, 1987–2020. Annual social-care expenditure among 5 771 603 adults aged ≥18 years identified from the Clinical Practice Research Datalink (CPRD) Gold and Aurum datasets linked to Hospital Episode Statistics (HES) and Office for National Statistics (ONS) data.

| Domain | N | Prevalence (%) | Mean £ per person in group | Total spend (£) | Spend per 100k adults (£) | Share of total spend (%) |
| --- | --- | --- | --- | --- | --- | --- |
| Personal Care | 418 578 | 7.25 | 8 736.00 | 3 656 697 408 | 63 356 704 | 84.26 |
| Mobility | 236 291 | 4.09 | 1 425.54 | 336 841 834 | 5 836 192 | 7.76 |
| Social Interaction | 488 282 | 8.46 | 708.70 | 346 044 855 | 5 995 645 | 7.97 |
| Multiple needs* | 342 874 | 5.94 | 9 076.06 | 3 111 945 821 | 53 918 224 | 71.71 |

(i) N: the number of people with non-zero annual spend in that domain. (ii) Prevalence (%): percentage of the total cohort with any recorded spend. (iii) Mean (£ per person in group): average annual cost among spenders (conditional on cost > 0). (iv) Total spend (£): sum of all individual annual costs in that domain. (v) Spend per 100 000 adults: calculated as (prevalence × mean × 100 000). (vi) Share of total spend (%): represents the proportion of total expenditure across the three main social care needs (Personal Care, Mobility, and Social Interaction), which together approximate 100%. (vii) Multiple needs:
*category includes individuals with spend in $\geq 2$ social care needs within the same person-year and is shown for context only; its totals are not additive with single-domain categories to avoid double counting.*

### Adjusted per-capita costs by age, sex, ethnicity, and deprivation

Adjusted per-capita annual costs for each social care need are shown in tables 3–6 and figure 2.

**Figure 2.**
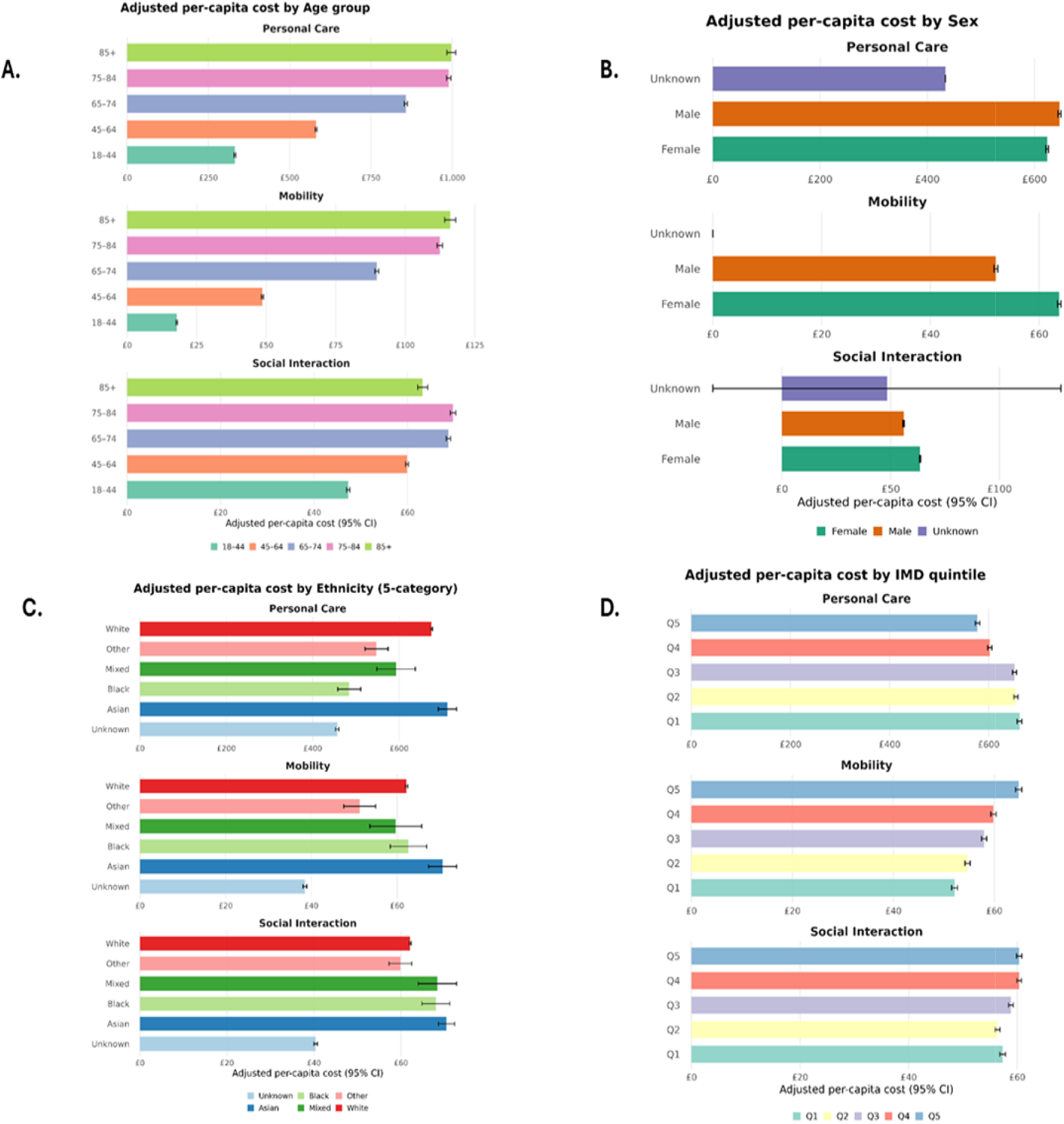
Adjusted annual per-capita social-care costs by demographic and socioeconomic subgroup among adults with multimorbidity in England, 1987–2020. Panels A–D show adjusted mean annual per-capita costs (£, 95% CI) for Personal Care, Mobility, and Social Interaction needs by age group (A), sex (B), ethnicity (C), and Index of Multiple Deprivation (IMD) quintile (D) among 5 771 603 adults aged ≥18 years with multimorbidity, using linked Clinical Practice Research Datalink (CPRD) Gold and Aurum data. Costs were directly standardised for sex, ethnicity, deprivation, and region, and include individuals with zero spend.

**Table 3.** Adjusted annual per-capita social care need costs by age group among adults with multimorbidity in England, 1987–2020. Adjusted mean annual per-capita costs (£) and 95% confidence intervals (CIs) for Personal Care, Mobility, and Social Interaction needs among 5,771,603 adults aged ≥18 years with multimorbidity, derived from linked Clinical Practice Research Datalink (CPRD) Gold and Aurum data. Estimates were directly standardised by sex, ethnicity, deprivation, and region, and include individuals with £0 spend to reflect population-level means.

| Domain | Stratum | Adj £ | 95% CI |
| --- | --- | --- | --- |
| **Mobility** | 18–44 | £17.9 | £17.6 to £18.1 |
|  | 45–64 | £48.6 | £48.3 to £49.0 |
|  | 65–74 | £89.7 | £89.1 to £90.4 |
|  | 75–84 | £112.4 | £111.5 to £113.4 |
|  | 85+ | £116.2 | £114.2 to £118.1 |
| **Personal Care** | 18–44 | £331.6 | £328.8 to £334.4 |
|  | 45–64 | £581.3 | £578.4 to £584.2 |
|  | 65–74 | £857.4 | £852.5 to £862.3 |
|  | 75–84 | £989.3 | £982.4 to £996.1 |
|  | 85+ | £997.0 | £983.5 to £1,010.5 |
| **Social Interaction** | 18–44 | £47.3 | £47.0 to £47.7 |
|  | 45–64 | £59.9 | £59.6 to £60.2 |
|  | 65–74 | £68.8 | £68.3 to £69.2 |
|  | 75–84 | £69.7 | £69.2 to £70.3 |
|  | 85+ | £63.3 | £62.2 to £64.3 |

**Table 4.** Adjusted annual per-capita social care need costs by sex among adults with multimorbidity in England, 1987–2020. Adjusted mean annual per-capita costs (£) and 95% confidence intervals (CIs) for Personal Care, Mobility, and Social Interaction needs among 5 771 603 adults aged ≥18 years with multimorbidity, derived from linked Clinical Practice Research Datalink (CPRD) Gold and Aurum data. Estimates were directly standardised for age, ethnicity, deprivation, and region, and include individuals with £0 spend. The “Unknown” category had very few observations and is presented for completeness only.

| **Domain** | **Stratum** | **Adj £** | **95% CI** |
| --- | --- | --- | --- |
| **Mobility** | Female | £63.7 | £63.4 to £64.0 |
|  | Male | £52.1 | £51.7 to £52.4 |
|  | Unknown | £0.0 | £0.0 to £0.0 |
| **Personal Care** | Female | £623.5 | £621.0 to £625.9 |
|  | Male | £646.6 | £643.8 to £649.4 |
|  | Unknown | £433.8 | £433.8 to £433.8 |
| **Social Interaction** | Female | £63.5 | £63.2 to £63.8 |
|  | Male | £55.9 | £55.7 to £56.2 |
|  | Unknown | £48.3 | -£31.8 to £128.3 |

**Table 5.** Adjusted annual per-capita social care need costs by ethnicity among adults with multimorbidity in England, 1987–2020. Adjusted mean annual per-capita costs (£, 95% CI) for Personal Care, Mobility, and Social Interaction needs by ethnicity among 5,771 603 adults aged ≥18 years with multimorbidity, derived from linked Clinical Practice Research Datalink (CPRD) Gold and Aurum data. Estimates were directly standardised for age, sex, deprivation, and region, and include individuals with zero spend. Costs were lowest among those with unknown ethnicity, reflecting incomplete recording rather than true differences in use.

| **Domain** | **Stratum** | **Adj £** | **95% CI** |
| --- | --- | --- | --- |
| **Mobility** | Asian | £70.6 | £67.3 to £73.9 |
|  | Black | £62.7 | £58.4 to £66.9 |
|  | Mixed | £59.7 | £53.7 to £65.7 |
|  | Other | £51.3 | £47.6 to £55.0 |
|  | White | £62.2 | £61.9 to £62.5 |
|  | Unknown | £38.5 | £38.0 to £38.9 |
| **Personal Care** | Asian | £712.0 | £690.9 to £733.2 |
|  | Black | £484.5 | £457.9 to £511.1 |
|  | Mixed | £593.1 | £548.7 to £637.5 |
|  | Other | £547.7 | £521.0 to £574.3 |
|  | White | £675.1 | £672.9 to £677.2 |
|  | Unknown | £456.5 | £452.7 to £460.3 |
| **Social Interaction** | Asian | £70.5 | £68.6 to £72.3 |
|  | Black | £68.0 | £64.9 to £71.2 |
|  | Mixed | £68.4 | £64.0 to £72.8 |
|  | Other | £59.9 | £57.2 to £62.5 |
|  | White | £62.1 | £61.9 to £62.3 |
|  | Unknown | £40.4 | £40.0 to £40.8 |

**Table 6.**
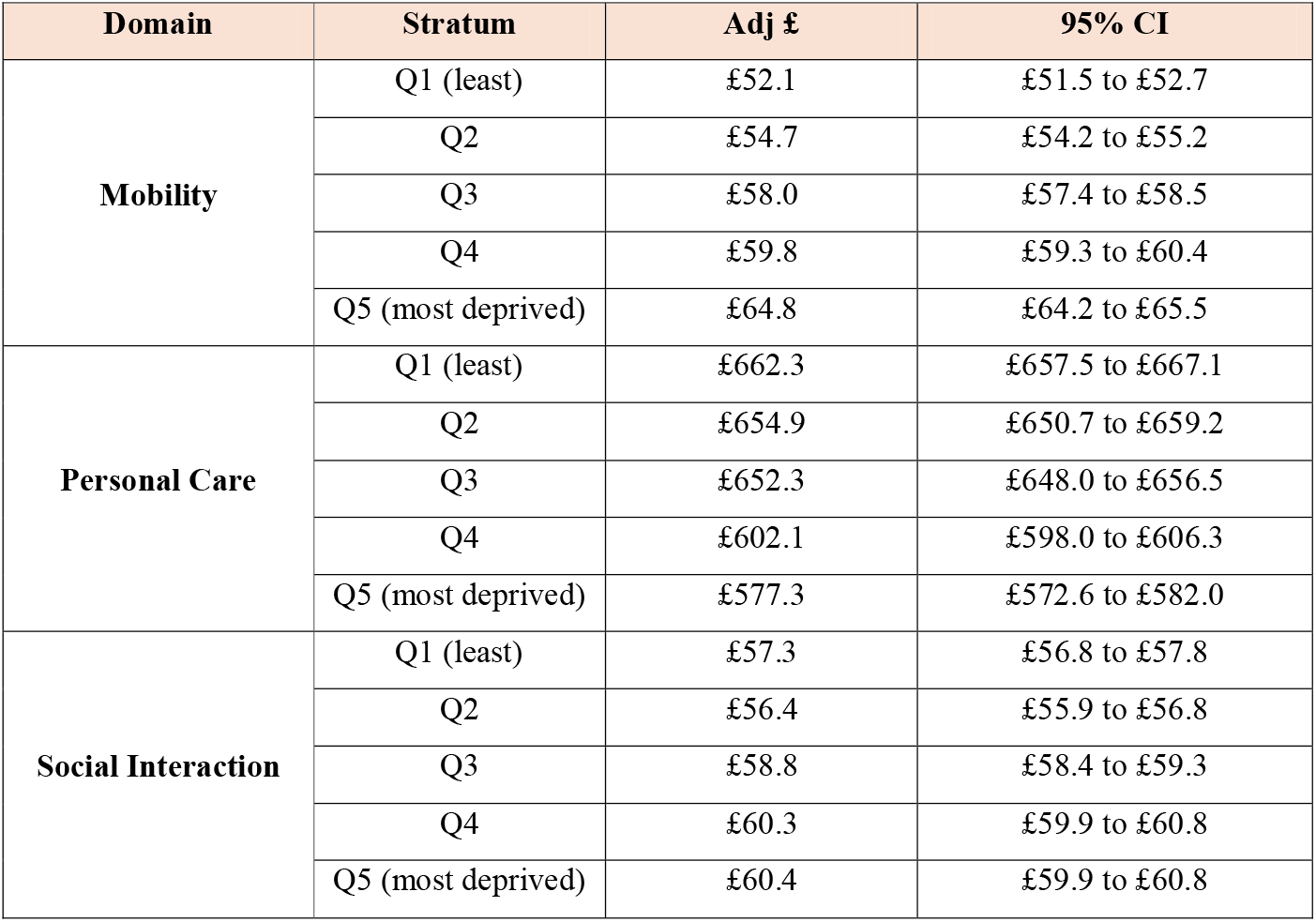
Adjusted annual per-capita social care need costs by socioeconomic deprivation among adults with multimorbidity in England, 1987–2020. Adjusted mean annual per-capita costs (£, 95% CI) for Personal Care, Mobility, and Social Interaction needs by Index of Multiple Deprivation (IMD) quintile among 5 771 603 adults aged ≥18 years with multimorbidity, using linked Clinical Practice Research Datalink (CPRD) Gold and Aurum data. Estimates were directly standardised for age, sex, ethnicity, and region, and include individuals with zero spend.

| **Domain** | **Stratum** | **Adj £** | **95% CI** |
| --- | --- | --- | --- |
| **Mobility** | Q1 (least) | £52.1 | £51.5 to £52.7 |
|  | Q2 | £54.7 | £54.2 to £55.2 |
|  | Q3 | £58.0 | £57.4 to £58.5 |
|  | Q4 | £59.8 | £59.3 to £60.4 |
|  | Q5 (most deprived) | £64.8 | £64.2 to £65.5 |
| **Personal Care** | Q1 (least) | £662.3 | £657.5 to £667.1 |
|  | Q2 | £654.9 | £650.7 to £659.2 |
|  | Q3 | £652.3 | £648.0 to £656.5 |
|  | Q4 | £602.1 | £598.0 to £606.3 |
|  | Q5 (most deprived) | £577.3 | £572.6 to £582.0 |
| **Social Interaction** | Q1 (least) | £57.3 | £56.8 to £57.8 |
|  | Q2 | £56.4 | £55.9 to £56.8 |
|  | Q3 | £58.8 | £58.4 to £59.3 |
|  | Q4 | £60.3 | £59.9 to £60.8 |
|  | Q5 (most deprived) | £60.4 | £59.9 to £60.8 |

Across age groups, all per-capita costs increased with age. Personal care costs increased from £331.6 (95% CI £328.8 to £334.4) among adults aged 18-44 years to £997 (£983.5 to £1,010.5) in those aged 85 years and older. Mobility needs cost from £17.9 (£17.6 to £18.1) in younger adults to £116.2 (£114.2 to £118.1) in the oldest age group. Social interaction costs from £47.3 (£47.0 to £47.7) in those aged 18-44 years to £69.7 (£69.2 to £70.3) in adults aged 75-84 years, before slightly declining to £63.3 (£62.2 to £64.3) in those aged 85 years or older.

By sex, women had higher adjusted per-capita costs for mobility (£63.7 [£63.4 to £64.0]) and social interaction (£63.5 [£63.2–£63.8]) compared with men (£52.1 [£51.7 to £52.4] and £55.9 [£55.7 to £56.2], respectively). Men, however, had marginally higher personal care costs (£646.6 [£643.8 to £649.4]) compared with women (£623.5 [£621.0 to £625.9]). Across ethnic groups, social care need costs were highest among Asian populations. For personal care needs, costs were highest among Asian adults (£712 [£690.0–£733.2]) and lowest among those with unknown ethnicity (£456.5 [£452.7 to £460.3]). Mobility needs costs ranged between £51.3 (£47.6–55) and £70.6 (£67.3–£73.9) across ethnic categories, with Asian adults showing the highest average. For social interaction needs, costs were broadly similar across groups, ranging from £59.9 (£57.2 to £62.5) in ‘Other’ ethnicities to £70.5 (£68.6 to £72.3) among Asian adults. Lower values in the Unknown category primarily reflect incomplete recording. By socioeconomic deprivation (IMD quintile), costs were higher with more deprivation for mobility and social interaction needs. Mobility needs costs increased from £52.1 (£51.5–£52.7) in the least deprived quintile to £64.8 (£64.2–£65.5) in the most deprived. Social interaction costs increased from £57.3 (£56.8–£57.8) in the least deprived to £60.4 (£59.9–£60.8) in the most deprived areas. In contrast, personal-care costs decreased with increasing deprivation, from £662.3 (£657.5–£667.1) in the least deprived to £577.3 (£572.6–£582) in the most deprived quintile.

## Discussion

In this study, we estimated the costs of social care need among individuals with multimorbidity in England using a large, population-based cohort derived from routinely collected electronic health records. Social care need was quantified through clinical and service-use indicators, and costs were assigned using standardised national unit estimates. Overall, we found that costs of social care increased with age, with those aged 85 and over incurring the highest costs, although, not for social interaction. Personal care need expenditure was by far the highest, £3.66 billion compared to £345 million for social interaction and £336.8 million for mobility. Differences in sex were observed, with increased costs for mobility and social interaction for females. Deprivation was associated with increased mobility and social interaction needs, whereas the least deprived had higher personal care costs. Lastly, overall social care need expenditure was highest within the Asian population, where costs were highest for social care needs and personal care needs. Taken together these findings elucidate the role of social determinants of health as predictors of social care expenditure.

### Comparison to existing literature

Results from this study support existing evidence that social care costs increase with age, particularly among those with multimorbidity [7]. However, we observed reduced costs for social interaction among individuals aged 85+, which may reflect unmet needs rather than lower demand. The Age UK (2024) report highlights that over two million older adults in England—especially those aged 80+ — face unmet social care needs due to complex health profiles and systemic limitations [25]. For example, our finding that personal care costs far exceeded that of mobility and social interaction costs, may reflect that social interaction needs are more difficult to assess and meet, than support with washing and other hygiene needs. Considering the WHO has recognised growing global rates of loneliness as a significant public health risk comparable to obesity [26]. In addition, workforce shortages and poor integration across primary, community, and social care services may contribute to delayed support, increased hospital admissions, and extended stays. Lower social interaction costs in this age group may therefore reflect reduced self-advocacy, inadequate service provision, or systemic neglect as opposed to less need.

We also found that females incurred higher costs than males, diverging from previous studies reporting no significant sex-based difference [27,28]. Client Level Data (2024) shows that women represent 56.1% of those receiving long-term support, with a notable overrepresentation in care homes [29]. This may be influenced by sociocultural norms around caregiving and age disparities between spouses, which can limit informal care provision [30]; males may be less willing to take on the caregiver role due to social-cultural stereotypingClick or tap here to enter text. [31]or, the tendency for women to marry men that are older than themselves means their partner may be willing but too frail themselves to provide the level of care needed for their spouse [32]

Our results revealed a non-linear relationship between deprivation and cost type. Individuals in the most deprived areas had higher costs for mobility and social interaction, while those in the least deprived incurred greater personal care costs—contrary to literature suggesting higher overall need in deprived populations [27,33]; So). Practical barriers such as digital exclusion, financial constraints, and stigma may limit access to care, resulting in lower recorded costs despite high need [34,35]. Systemic inequities, including fewer GPs per patient and shorter consultations in deprived areas [36], may further contribute to unmet needs.

Lastly, we found that Asian individuals had higher social care costs, which contrasts with previous reports indicating lower uptake among ethnic minorities. While cultural preferences for informal care and barriers such as stigma and language have historically limited access[31,37], our results may reflect a shift in attitudes toward formal care within Asian communities, consistent with recent increases in service use [38].

### Strengths and limitations

To our knowledge, this study is among the first to use individual-level, longitudinal data from a nationally representative sample to quantify the economic burden of social care need in people with multimorbidity at scale in England. Leveraging linked primary care and administrative datasets provides an opportunity to measure individual-level social care costs at national scale. The large cohort size of 800 723 and long follow-up enabled detailed estimation of costs across sociodemographic groups and clinical subpopulations, providing insights not possible with survey-based or ecological data. All costs were standardised to 2019-2020 PSSRU estimates ensuring comparability and alignment with recent nationally available costs data. Furthermore, the inclusion of individuals across the age spectrum (≥18 years), both sexes, diverse ethnic groups, multiple deprivation quintiles and a wide range of multimorbidity and social care needs ensured generalisability to the wider English population. The use of routinely collected electronic health records linked with national cost data avoids the biases inherent in self-report approaches, allowing for standardised cost attribution across the cohort.

At the same time, several important limitations should be noted. Most critically, this study excluded approximately 6.2 million individuals (88% of the multimorbidity cohort) who had no recorded social care costs. This means our analysis measures service utilisation among those accessing formal care, not population-level social care need. This fundamentally limits our ability to draw conclusions about equity. Lower costs observed in some groups could reflect systematic barriers to accessing services— our study design cannot distinguish between these possibilities. The true burden of unmet need in these populations remains unknown and may be substantial.

Additionally, social care need was operationalised through proxy indicators derived from healthcare datasets, which may not fully capture the breadth of need or provision. In particular, informal and unpaid care, privately funded services, and unmet need are not reflected in our cost estimates. While we adjusted for a range of demographic and clinical characteristics, residual confounding may remain, especially for unmeasured factors such as functional status, cognitive impairment, or the availability of family support. Some indicators may be more consistently recorded than others, introducing potential bias. Data completeness also varied, and exclusion of individuals with missing values may limit representativeness in certain subgroups. Finally, cost estimates relied on assumptions and proxies where direct PSSRU unit costs were unavailable, which could introduce measurement error, though these assumptions were applied systematically across the cohort.

### Implications and conclusions

Multimorbidity substantially increases social care need costs in England, but these costs are not distributed evenly. The mixed patters of expenditure among deprived and older age groups may suggest inequities in access, provision, or uptake of services. Multimorbidity burden is predicted to increase in coming years along with the costs that come with its care. Social care thus constitutes a fundamental aspect of care planning. Policy responses must recognise that healthcare costs capture only part of the multimorbidity burden. Social care funding has declined in real terms over the past decade, leaving local authorities and unpaid carers to absorb much of the demand. Without investment and reform, rising multimorbidity will intensify pressures on already stretched social care systems. To address this challenge, health and social care planning should be integrated, with explicit recognition of the costs and inequalities associated with multimorbidity. National funding models must account for both health and social care to avoid systematic underestimation of need. Targeted strategies are required to improve equity of access, alongside support for carers and community-based services. Strengthening the routine recording of social care in administrative datasets, and linking it with healthcare information, will be essential for monitoring inequalities and guiding resource allocation.

## Data Availability

The data that support the findings of this study are available from CPRD but restrictions apply to the availability of these data, which were used under license for the current study, and so are not publicly available. Data are however available from the authors upon reasonable request and with permission of CPRD.

https://www.cprd.com/access-data

## Declarations

### Ethics approval and consent to participate

Ethical approval for this study was granted by the University of Southampton Faculty of Medicine Research Committee (reference: 67953). The study was also approved by the Independent Scientific Advisory Committee (ISAC) for the Clinical Practice Research Datalink (CPRD) (protocol number: 21_001667). All methods were performed in accordance with the relevant guidelines and regulations, including those set out by the CPRD and the Declaration of Helsinki.

CPRD collects anonymised primary care data from practices that have consented to contribute. Patients can opt out of data sharing, and no data are collected for those who have opted out. As all data used in this study were de-identified and routinely collected for health care delivery, informed consent from individual patients was not required.

### Authors’ contributions

HDM conceptualised the study and wrote the first draft of the manuscript. TI and RV conducted the data analysis, with input from AJ, LS, YY and MH. TI, LS, and HDM drafted and revised the manuscript, with additional review and formatting by YY, MH and AJ. YY, AJ, and MH reviewed the R code and contributed to revisions. All authors had full access to all data in the study and reviewed and approved the final version of the manuscript. HDM had final responsibility for the decision to submit for publication.

## List of Abbreviations

ADL: Activities of Daily Living
BMC: BioMed Central
CI: Confidence Interval
CPRD: Clinical Practice Research Datalink
HDM: Hajira Dambha-Miller
IADL: Instrumental Activities of Daily Living
ICD-10: International Classification of Diseases, 10th Revision
IMD: Index of Multiple Deprivation
ISAC: Independent Scientific Advisory Committee
LTC: Long-Term Condition
NIHR: National Institute for Health and Care Research
NHS: National Health Service
OR: Odds Ratio
PSSRU: Personal Social Services Research Unit
Q: Quintile/Quartile
SD: Standard Deviation
SNOMED: Systematized Nomenclature of Medicine
UK: United Kingdom

