## Supplementary table 2 for "The Cost of Social Care Need in People with Multimorbidity: A Population-Based Cohort Study of 5.7 million Individuals"

**Supplementary Material 1**

**Table S1. Cohort selection and exclusions for adults with multimorbidity (**≥2 long-term conditions), 1987-2020

Cohort selection flow from the Clinical Practice Research Datalink (CPRD) Gold and Aurum databases, linked to Hospital Episode Statistics and covering the period 1987-2020. The table presents the sequential application of inclusion criteria among adults with multimorbidity (defined as having ≥2 long-term conditions). For each step, the total number of individuals retained, and the number and percentage excluded, are shown.

| **Step** | **N** | **Dropped (N)** | **Dropped (%)** |
| --- | --- | --- | --- |
| 0) CPRD Gold and AURUM with =< 2 long-term conditions | 7290716 |  |  |
| 1) In study window (index_date present) | 5856089 | 1434627 | 19.68 |
| 2) Adults (age_index > = 18) | 5771603 | 84486 | 1.44 |

**Table S2.** List of Long-Term Conditions (LTC) identified in the Clinical Practice Research Datalink (CPRD) Database, mapped against the original 59 LTC conditions defined through national consensus. Some conditions have been grouped based on their availability in the CPRD Database.

|  | **Long-Term Conditions in the Original 59 Conditions** | **Long-Term Conditions Present in The Study** |
| --- | --- | --- |
| 1 | Addison’s Disease | Yes |
| 2 | Anaemia | Yes  Present as Pernicious anaemia |
| 3 | Anxiety | Yes |
| 4 | Depression | Yes |
| 5 | Congenital Heart Disease | Yes |
| 6 | Chromosomal Abnormality | Yes |
| 7 | Bipolar Disorder | Yes |
| 8 | Schizophrenia | Yes |
| 9 | Chronic Liver Disease | Yes |
| 10 | Alcohol-related Liver Disease | Merged and renamed as: Chronic Liver Disease and Alcoholic Liver Disease |
| 11 | Aortic Aneurysm | Yes |
| 12 | Arrhythmia | Yes |
| 13 | Asthma | Yes |
| 14 | Autism | Yes |
| 15 | Bronchiectasis | Yes |
| 16 | Cancer | Yes |
| 17 | Cerebral Benign Tumours | No |
| 18 | Chronic Back Pain | No |
| 19 | Chronic Lyme Disease | Yes |
| 20 | Chronic Pancreatitis | Yes |
| 21 | Chronic Kidney Disease stage 3-5 | Yes |
| 22 | Chronic Pain | Yes |
| 23 | Chronic Obstructive Pulmonary Disease | Yes |
| 24 | Chronic Urinary Tract Infections | Yes |
| 25 | Connective Tissue Disease | Yes |
| 26 | Coronary Heart Disease | Yes |
| 27 | Heart Failure | Yes |
| 28 | Hypertension | Yes |
| 29 | Cystic Fibrosis | Yes |
| 30 | Dementia | Yes |
| 31 | Diabetes | Yes |
| 32 | Drug or Alcohol Misuse | Yes |
| 33 | Eating Disorders | Yes |
| 34 | Endometriosis Adenomyosis | Yes |
| 35 | Epilepsy | Yes |
| 36 | Gout | Yes |
| 37 | Hearing Loss | Yes |
| 38 | Hemi/Para/Quadriplegia | No |
| 39 | HIV/AIDS | Yes |
| 40 | Inflammatory Bowel Disease | Yes |
| 41 | Long COVID | Yes  Present as Post-Acute COVID 19 |
| 42 | Meniere’s Disease | Yes |
| 43 | Multiple Sclerosis | Yes |
| 44 | Musculoskeletal Injury | Yes |
| 45 | Osteoarthritis | Yes |
| 46 | Osteoporosis | Yes |
| 47 | Parkinson’s Disease | Yes |
| 48 | Peptic Ulcer Disease | Yes |
| 49 | Peripheral Neuropathy | Yes |
| 50 | Peripheral Vascular Disease | Yes |
| 51 | Post-Traumatic Stress Disorder | Yes |
| 52 | Sickle Cell Disease | No |
| 53 | Stroke | Yes |
| 54 | Thyroid Disease | Yes |
| 55 | Transient Ischemic Attack | Yes |
| 56 | Tuberculosis | Yes |
| 57 | Valvular Diseases | Yes  Present as Heart Valvular Disorder |
| 58 | Venous Thromboembolism | Yes |
| 59 | Visual Impairment | Yes |

**Table S3. List of long-term conditions and corresponding clinical codes used to define multimorbidity, 1987-2020.**

List of all long-term conditions included in the analysis, with corresponding Read and SNOMED CT codes used to identify diagnoses in the Clinical Practice Research Datalink (CPRD) Gold and Aurum databases linked to Hospital Episode Statistics (HES). Multimorbidity was defined as the presence of ≥ 2 of these long-term conditions on or before the study index date (1 January 1987).

Table in healtheconomics Supplemental file 2

**Table S4. Summary of activities consisting of Social Care Needs (SCN)**

**Activities of Daily Living (ADL)**

Received professional care or support for ADL

Difficulty handling food

Difficulty with toiletry

Difficulty with gardening

Difficulty taking a shower

Difficulty using stairs

Difficulty with physical activities

Difficulty with bathing

Difficulty using appliance

Difficulty travelling

Difficulty with personal care

Difficulty managing medication

Difficulty performing clerical activities

Difficulty cleaning home

Difficulty in walking

Difficulty performing intellectual activities

Difficulty using non-verbal and verbal communication

Difficulty hearing

Difficulty understanding verbal and written language

Difficulty speaking

**Social Network Needs**

Difficulty with communication

Support provided professionally to assist social networking and social engagement

Referral to social prescriber

Difficulty travelling to take part in social network activities

Difficulty with social participation

**Mobility Needs**

Able to mobilise using mobility aid

Difficulty with mobility

Do not mobilise using aid

Difficulty mobilising indoor

Difficulty managing stairs

Mobilise using aid

Mobility assessed using the modified elderly mobility scale

Mobility assessed using the shuttle walking test

Difficulty mobilising outdoor

Difficulty walking

**Table S5. Definition of social care need domains and corresponding Clinical Practice Research Datalink (CPRD) variables, 1987-2020.**

List of all social care need domains and the corresponding CPRD Gold and Aurum variables, Read and Medcodes used for classification. Variables were grouped into eight validated domains representing distinct aspects of social vulnerability and dependency: Hearing disorders, Financial challenges, Social isolation, Mobility impairments, Bereavement, Learning disabilities, Residential needs, and Limitations in activities of daily living (ADL). All variables were standardised across CPRD Gold and Aurum and mapped to relevant Medcodes and Read codes. Each domain was coded as binary (1 = presence of need; 0 = absence) at the individual level, based on any recorded indicator during the study period (1987–2020).

SOCIAL-NEED-CODES Healtheconomics Supplemental file 3

**Table S6. Source of unit cost estimates for social care needs.**

All unit cost data for social care needs were extracted from the Personal Social Services Research Unit (PSSRU) “Unit Costs of Health and Social Care 2020” report. The full dataset is publically available online at [Unit Costs of Health and Social Care 2020 | PSSRU](https://www.pssru.ac.uk/project-pages/unit-costs/unit-costs-2020/) .

**Table S7. Costing methodology for social care needs applied in this study.**

SocialCare_Costing healtheconomics Supplemental file 4

**Table S8. Overlap of social care needs among adults with multimorbidity (≥ 2 long-term conditions), 1987–2020**

Overlap matrix showing unique combinations of Personal Care, Mobility, and Social Interaction needs among 5 771 603 adults aged ≥ 18 years with multimorbidity, using linked Clinical Practice Research Datalink (CPRD) Gold and Aurum data (1987–2020). Each row represents a mutually exclusive group, where “1” indicates the presence and “0” the absence of the social care need. Counts (N) refer to the number of individuals in each category.

| **Personal care needs** | **Mobility needs** | **Social interaction needs** | **N** |
| --- | --- | --- | --- |
| 0 | 0 | 0 | 4971294 |
| 1 | 0 | 1 | 286579 |
| 0 | 0 | 1 | 201812 |
| 0 | 1 | 0 | 161361 |
| 1 | 0 | 0 | 75574 |
| 1 | 1 | 1 | 37402 |
| 1 | 1 | 0 | 19023 |
| 0 | 1 | 1 | 18558 |

**Table S9. Costs for social care needs used in the cost assignment.**

List of annual equivalent unit costs (£, 2020 prices) applied to social care needs in the cost-assignment framework. Costs were extracted from Curtis L & Burns A. (2020) Unit Costs of Health and Social Care 2020, Personal Social Services Research Unit (PSSRU), University of Kent, Canterbury [Unit Costs of Health and Social Care 2020 | PSSRU](https://www.pssru.ac.uk/project-pages/unit-costs/unit-costs-2020/) . PSSRU Ref refers to the source codes used in PSSRU cost sheet identifiers. London uplifts (+14% for private nursing homes and +18% for private residential care) were applied where indicated. All values represent annual per-person costs unless otherwise specified.

| **PSSRU Ref** | **Sector** | **Residential Status** | **Sub-Group** | **Age bracket** | **Frequency** | **Weekly Costs** | **Annual costs** | **Notes** |
| --- | --- | --- | --- | --- | --- | --- | --- | --- |
| ADL Needs | | | | | | | | |
| 1.1 | Private (for-profit) | Nursing home | Older people | 65+ | per week | 907 | 47327.26 | A-E weekly (incl. living expenses & external services). Occupancy ~91%. London +14%. |
| 1.3 | Local authority | Residential care | Older people | 65+ | per week | 1291 | 67364.38 | A-I weekly (capital, LA exp, living & external). Occupancy 92.6%. |
| 4.3 | Mixed | Residential care | Learning disability | 18-64 | per week | 1578 | 82340.04 | ASC-FR 2018/19 median weekly cost uprated to 2019/20. |
| 4.4.2 | Specialist provider | Residential care | Autism & complex needs | 18-64 | per year | 100755 | 100755 | Annual cost per client excl. day care; incl. day care: £127,879/yr. |
| 12.5 | Independent sector | Home care (domiciliary) | Adults (Social services) | All adults | per hour | 24 | 12355 | Average cost across 7 h/week and 12.8h/week |
| 19 | For-profit | Residential care | Physical disability | All adults | per week (benchmark) | 433 | 22593.94 | Midpoint between min & max weekly fee. |
| 19 | Non-profit | Nursing home | Dementia | All adults | per week (benchmark) | 1137 | 59328.66 | Midpoint between min & max weekly fee. |
| 19 | Non-profit | Residential care | Dementia | All adults | per week (benchmark) | 729 | 38039.22 | Midpoint between min & max weekly fee. |
| 19 | Non-profit | Nursing home | Learning disability | All adults | per week (benchmark) | 1317 | 68721.06 | Midpoint between min & max weekly fee. |
| Mobility Needs | | | | | | | | |
| 19 | Non-profit | Nursing home | Physical disability | All adults | per week (benchmark) | 1396 | 72843.28 | Midpoint between min & max weekly fee. |
| 5.1 | Local authority | Residential care | Physical support | 18-64 | per resident-week | 1272 | 66372.96 | A-E weekly (incl. capital via A-C and personal living expenses). Occupancy assumed 100%. |
| 5.2 | Voluntary/Private | Residential care | Physical support | 18-64 | per resident-week | 1000 | 52180 | A-E weekly (incl. personal living expenses). Occupancy assumed 100%. |
| 5.3 | Local authority | Day care | Physical support | 18-64 | per attendance | 100 | 14040 | Assume 2.7 attendances/week x— 52 weeks; ~4.8h per attendance. £21/hour available. |
| 5.3 | Discount by 3.5 for over 60 | Day care | Physical support | over 60 | per attendance | 96.5 | 9126 | Assume 2.7 attendances/week x— 52 weeks; ~4.8h per attendance. £21/hour available. |
| 7.2 | NHS | Wheelchair service | Mobility equipment | All adults | per chair-year | 103 | 103 | Capital annuity ~£71 + maintenance £32; 5-year life, 3.5% discount. |
| 7.2 | NHS | Wheelchair service | Mobility equipment | All adults | per chair-year | 209 | 209 | Capital annuity ~£178 + maintenance ~£32; 5-year life, 3.5% discount. |
| 7.2 | NHS | Wheelchair service | Mobility equipment | All adults | per chair-year | 481 | 481 | Capital annuity ~£355 + maintenance £126; 5-year life, 3.5% discount. |
| Social Network Needs | | | | | | | | |
| 1.4 | Local authority | Day care | Older people | 65+ | per attendance | 64 | 8320 | Assume 2.5 attendances/week x— 52 weeks. Also available: £14/hour; £49 per 3.5h session. |
| 4.1 | Local authority | Day care | Learning disability | 18-64 | per attendance | 72 | 17971.2 | Assume 4.8 attendances/week x— 52 weeks; ~4h per attendance. Also available: £15.50/hour; £46.50 per 3.5h session. |
| 2.3 | Local authority | Day care | Adults with mental health support needs | 18+ | per attendance | 39 | 6084 | Assume 3 attendances/week x— 52 weeks; also £9.48/hour; £33 per 3.5h session. |
| 11.7 | LA/Voluntary/Independent | Community outreach | Adults (community engagement) | 18+ | per hour | 25 | 3913.5 | Annual = hourly x 3 hours/week x— 52.18. Example: 1h/wk=£1,300; 2h/wk=£2,600. |
| 19 | Non-profit | Nursing home | Mental health | All adults | per week (benchmark) | 790 | 41222.2 | Midpoint between min & max weekly fee. |
| 19 | Non-profit | Residential care | Mental health | All adults | per week (benchmark) | 716 | 37360.88 | Midpoint between min & max weekly fee. |
