## Supplementary table 3 for "The Cost of Social Care Need in People with Multimorbidity: A Population-Based Cohort Study of 5.7 million Individuals"

### Supplementary Methods

#### Cost mapping and assignment

Mappings were based on provider type, setting, client group, age band, unit type, annual equivalent cost (GBP), and London uplifts. London uplifts were +14% for private nursing homes (code 1.1*) and +18% for private residential care (code 1.2); no uplift applied to local-authority residential care (code 1.3*).

#### Deterministic cost assignment

To assign costs to individuals, a closest-match engine was implemented. For each combination of setting, client group, and age band, each cost record received an age-match score and a client-group score valued as: 2 for an exact match, 1 for a compatible match (e.g., age 18–64 matched to “All adults”, or age ≥65 matched to “65+” or “All adults”), and 0.5 when unspecified. A setting score of 1 was given when the cost setting overlapped the CPRD setting. The total score was computed as 1.5 × age + 1.5 × client + 1.0 × setting. The row with the highest total score was selected, by source code. When a London uplift applied, the annual price was multiplied by (1 + uplift) for residents identified as living in London. Costs were treated as annual equivalents per person-year unless the tariff referred to per-episode (e.g., reablement) or per-hour (e.g., peer support). Hourly tariffs were converted to yearly equivalents only when default weekly hours were specified.

#### Personal-care costing

Personal-care costing followed a hierarchical trigger set to ensure one personal-care package per person-year, avoiding overlap between residential per-week tariffs and community hourly tariffs. Individuals ≥65 years with nursing-home status were assigned a nursing-home tariff for older people via the closest-match engine, preferentially selecting private nursing-home 1.1 (+14% London uplift) where applicable, or the dementia benchmark line when dementia was recorded and no more specific line existed. Individuals ≥65 years with residential-care status were assigned a residential-care tariff (private 1.2 +18% London uplift or local-authority 1.3 if closer). Individuals 18–64 with a learning-disability flag were assigned supported living (4.3.2) when appropriate, or residential (4.3) otherwise. Individuals 18–64 with autism were assigned supported living (4.4.1) or residential autism (4.4.2) as matched. If dementia was present and none of the above applied, the nursing-home dementia benchmark (code 19*) was used. When no residential trigger was present but personal-care need was indicated (pc_any = 1), a community home-care package (11.5) was assigned using the 7 h/week assumption (12.8 h/week retained for sensitivity analyses). Each person received one personal-care setting, an annual cost, and the catalogue code used in assignment.

#### Mobility costing

Mobility costing included three primary components, with higher-risk overlaps for sensitivity analyses: (1) Wheelchair and mobility equipment: triggered by a wheelchair/aids indicator; intensity tiers—basic £103, mid £209, high £481 per year, determined by total mobility-item count (thresholds at 3 and 5 items). (2) Home adaptations: triggered by functional barriers indoors/outdoors/on stairs or walking; annualised installation-year bands £660 (one barrier), £1,426 (two), £3,264 (≥3). (3) Reablement: one episode per year when reduced mobility or a mobility assessment was recorded, costed at £1,728 when a Waterlow flag was present and £1,484 otherwise. Day services per attendance, residential “physical support 18–64 per resident-week,” and inpatient neuro-rehabilitation per occupied bed day were excluded in the primary analysis to avoid double counting with residential tariffs; they were included only in sensitivity analyses. The mobility annual cost per person equalled the sum of active components, with matched codes retained for audit.

#### Social-interaction costing

Social-interaction costing combined day-care, social-prescribing, and peer-support components. Individuals ≥65 years with difficulty in social participation were assigned local-authority older-people day care (annualised at 2.5 attendances/week). Individuals 18–64 with social-participation difficulty and a learning-disability flag were assigned learning-disability day care (4.8 attendances/week). Adults with mental-health needs and social-participation difficulty were assigned adult mental-health day care using local-authority or private/voluntary lines. Social prescribing was assigned when a social-prescriber referral was recorded; the middle unit price (£398) among £177–£570 was used as primary. Peer support was assigned at 12 hours/year × £28/hour; £39/hour was used in sensitivity analyses. Community outreach was priced only when hours were recorded. The social-interaction annual cost per person equalled the sum of active components.

#### Avoiding double counting

Double counting was prevented through hierarchical rules: (1) Residential personal-care tariffs took precedence over mobility and social-interaction items and were not combined with community hourly tariffs. (2) Only one residential personal-care category was assigned per person. (3) Mobility “physical support per resident-week” tariffs were disabled whenever a residential personal-care tariff was assigned.

*codes refer to codes available in cost sheets provided by *Unit Costs of Health and Social Care 2020*. Personal Social Services Research Unit (PSSRU), University of Kent, Canterbury [Unit Costs of Health and Social Care 2020 | PSSRU](https://www.pssru.ac.uk/project-pages/unit-costs/unit-costs-2020/)
